# Deep-learning-based automatic detection of pulmonary nodules from chest radiographs

**DOI:** 10.1101/2022.06.21.22276691

**Authors:** Pranav Ajmera, Richa Pant, Jitesh Seth, Suraj Ghuwalewala, Sahil Kathuria, Snehal Rathi, Sonali Patil, Manaswani Edara, Mukul Saini, Preeti Raj, Vinay Duddalwar, VM Kulkarni, Parag Patil, Viraj Kulkarni, Amit Kharat

## Abstract

**Objective:** To assess a deep learning-based artificial intelligence model for the detection of pulmonary nodules on chest radiographs and to compare its performance with board-certified human readers.

**Methods:** For this retrospective study, 308 chest radiographs were obtained between January 2019 to December 2021 from a tertiary care hospital. All radiographs were analyzed using a deep learning AI model called DxNodule AI Screen. Two expert board-certified radiologists established the ground truth, and 11 test readers independently reviewed all radiographs in two sessions (unaided and AI-aided mode) with a washout period of one month.

**Results:** The standalone model had an AUROC of 0.905 [0.87, 0.94] in detecting pulmonary nodules. The mean AUROC across the 11 readers improved from 0.798 [0.74, 0.86] for unaided interpretation to 0.846 [0.82, 0.880] for AI-aided interpretation. With DxNodule AI Screen, readers were able to identify nodules at the correct locations, which they otherwise missed. The mean specificity, accuracy, PPV, and NPV of the readers improved significantly from 0.87 [0.78, 0.96], 0.78 [0.72, 0.84], 0.77 [0.65, 0.88], and 0.86 [0.81, 0.90] in the unaided session to 0.89 [0.82, 0.96], 0.83 [0.80, 0.85], 0.82 [0.73, 0.9], and 0.89 [0.86, 0.92], respectively in the aided session.

**Conclusion:** DxNodule AI Screen outperformed human readers in nodule detection performance on chest radiographs, and enhanced human readers’ performances when used as an aid.

## INTRODUCTION

Chest radiography is the most frequently advised and often the first-line imaging investigation for the diagnosis of suspected respiratory disorders, including lung cancer. This is due to the ease of procedure, wide-scale availability, accessibility, and lower radiation exposure as compared to chest CT scans. The ease of the procedure ensures early identification of lesions, allowing the timely assessment and management of the disease^1^. However, reports suggest that 19-26% of the lung nodules apparent on the chest radiographs are overlooked at their first readings ^2,3^. This happens because radiologists and clinicians may fail to identify a nodule when looking for more common disease manifestations ^4^. However, a nodule can have a range of etiological causes and if the etiology is malignant, the patient is likely to present later with a more widely spread neoplasia, negatively influencing both patient morbidity and mortality. Missed lung cancer is a subject of concern among radiologists and is a serious medicolegal issue. In fact, undetected pulmonary lesions are amongst the most common causes of malpractice issues ^5^.

As per GLOBOCAN-2020, lung cancer remains the most common cause of mortality due to cancer, accounting for nearly 1.8 million deaths and 2.1milion new cases each year ^6^. Multiple retrospective studies have shown that, while chest radiography is the first line and most commonly advised imaging modality for detecting a pulmonary nodule/ lung cancer, it is also the most common cause of missed lesion identification, accounting for nearly 90% of all missed lung cancers. On the other hand, CT examinations account for only 5% of missed nodules/ lung cancer ^7^. Amongst the multiple contributory causes of a missed diagnosis, the most common cause is observer error ^8^. The experience of the reporting radiologist also has a near-direct correlation with the likelihood of identifying a nodule on a chest radiograph ^9^. The possible causes behind these overlooked lesions are two-pronged: an error in detection and an error in recognition. These lesions are missed by radiologists due to a lack of adequate skill set, an improper chest radiograph scanning pattern, or misclassification of the radiograph as another pathology ^4^. Therefore, many computer-aided diagnostic techniques have been proposed in recent times, resulting in improvement in reader performances. Artificial intelligence (AI) is demonstrating rapid improvements and achievements in the field of diagnostic imaging ^10,11^. A reliable AI system can assist human readers in diagnosing cases more quickly and accurately. The AI-powered technology has the potential to identify pulmonary nodules at an early stage, with detection capability comparable to an expert reader. The use of a large training dataset exposes the AI model to the entire spectrum of the disease, allowing it to recognize nodules on chest radiographs in clinical practice. By screening and identifying each radiograph, such a model can improve the performance of reviewing radiologists and save their time, allowing them to do more targeted and speedy reporting ^12^.

Herein, we evaluate the performance of a deep learning model in detecting pulmonary nodules on chest radiographs and compare its performance with that of consultant human readers with varied experience in diagnosing the disease.

## MATERIALS AND METHODS

The study was approved by the institutional review board of Dr. D.Y. Patil hospital and research center as a part of its ethical committee meeting (DYPV/EC/740/2021). The Institutional Review Board (IRB) waived the need for separate consent from individual patients in view of the retrospective and observational nature of the study. The data was extracted and utilized in a Health Insurance Portability and Accountability Act (HIPAA) compliant format while ensuring complete anonymization of the patient data from individual radiographs.

### Data Acquisition

The study included 308 frontal chest radiographs (220 PA and 88 AP) from 308 adult patients from OPD as well as IPD settings. The radiographs were acquired on multiple machines of different milliamperage (mA). These included multiple computed radiography (CR) systems: SIEMENS 500mA HELIOPHOS-D, SIEMENS 100mA GENIUS-100R, SIEMENS 300mA MULTIPHOS-15R, and a 600mA digital radiography (DR) system: the SIEMENS MULTI SELECT DR. For the CR systems, AGFA 14×17 inch plate was used, and for the DR system, SIEMENS 14×17 inch detector was used.

### Data Retrieval

For data collection, two board-eligible radiology residents (S1, S2) assessed and identified all chest radiographs that were reported as “suspicious pulmonary nodule/ suggestive of the nodule/ suggestive of the pulmonary nodule/ indicative of pulmonary nodule” from the institute’s Picture Archival and Communication System (PACS). The search was performed for the frontal chest radiography performed between January 2019 and December 2021 and yielded 324 chest radiographs. S1 and S2 assessed all chest radiographs for their quality and fit within the inclusion/exclusion criteria. Poor quality radiographs that were end-expiratory, oblique, underexposed, overexposed, or exhibited motion blur, were excluded from the study. Radiographs that included a partial field of interest and radiographs with artifacts and degradations that interfered with normal reading were also excluded from the study. All radiographs included in the study had to meet additional image quality criteria, such as the Digital Imaging and Communications in Medicine (DICOM) image format standard, and coverage of the entire lung field. Following the implementation of the exclusion and inclusion criteria, 308 radiographs were qualified to be included in the clinical validation study.

### Establishing the Ground-truth

To establish the ground truth, three experienced board-certified radiologists G1 (with 23 years of experience), G2 (with 7 years of experience), and G3 (with 5 years of experience) assessed all radiographs. All three radiologists labeled the presence or absence of nodules in a consensus reading fashion. The radiologists establishing the ground truth did not have the access to AI-generated results to avoid any bias.

### Defining the Target Disease

As per the Nomenclature Committee of the Fleischner Society, a pulmonary nodule is defined as an ovoid to rounded hyperdense radiopacity within the pulmonary interstitium, with an absence of air-bronchogram. Pulmonary nodules are mostly well-demarcated but can sometimes be poorly delineated. Their size is typically less than 3cm ^13^.

### AI Model development

For preprocessing, the X-Ray DICOM images were converted to 8-bit grayscale images of size 768 × 768, and the pixel values were normalized to lie between 0 and 1. Various image augmentation techniques were used during model training. In each epoch, half of the input images were either flipped horizontally or had their brightness or contrast modified. The processed images were then used for training the model. The deep learning model architecture was an ensemble of two Feature Pyramid Networks (FPN1 and FPN2), each having an Xception encoder. The difference between the two FPNs was the loss function. While FPN1 was trained using both Binary CrossEntropy (BCE) and Dice loss equally, FPN2 used 0.9 BCE + 0.1 Dice loss (see Fig. 1). Both models were trained using the “Adam” optimizer and a learning rate of 0.001 (with “ReduceLROnPlateau” scheduler) was used to regulate the learning while training ^14^.

**Figure 1:**
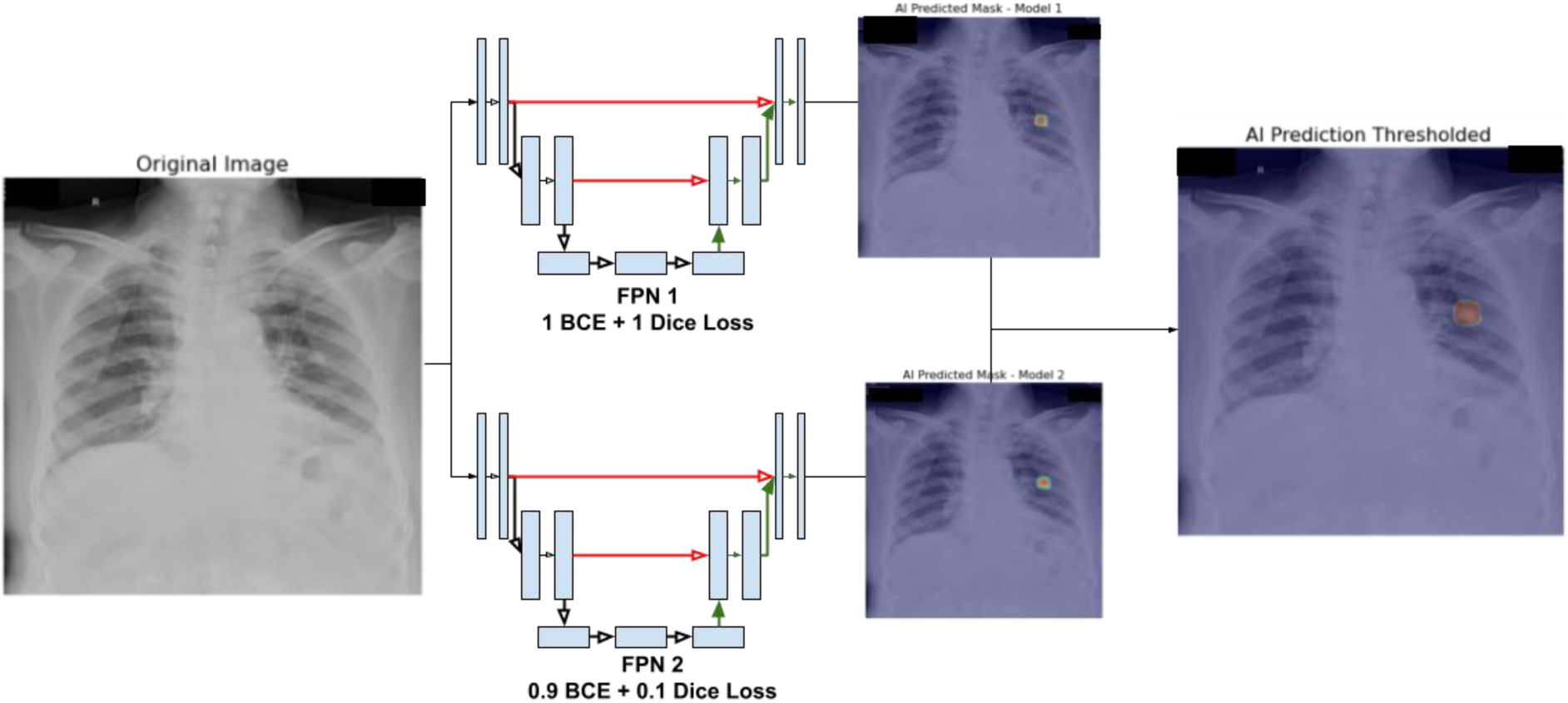
A schematic representation of the pulmonary nodule detection workflow.

### Observer performance study

To assess the performance of a DL model, and establish its clinical utility, it is necessary to evaluate the model performance against the performance of multiple readers. It is also important to evaluate the differences in the performance of readers when the DL output is provided as an aid to them. For the observer performance study, chest radiographs were anonymized and randomized across readers and reading sessions. The radiologists who determined the ground truth did not participate as observers. We compared the performance of 11 readers without and with the aid of DxNodule AI Screen. Readers participating in the study included a physician with 3 years of experience in general medicine and 2 years of experience as a medical officer; an emergency medicine consultant with 4 years of dedicated field experience; a pulmonologist with 3 years of experience and a preceding 2 years of experience as a general practitioner; an anesthesiologist with 3 years of experience who had been posted as an ICU in-charge for over a year; a radiologist who had 5 years of experience but had subspecialty training in cross-sectional and musculoskeletal imaging; an X-ray technologist; and 5 resident radiologists.

Observers were blinded to patient information, reference standard, final diagnosis, and the ratio of radiographs with and without nodules. All readers reviewed radiographs in two sessions (without and with DxNodule AI Screen outputs) and recorded their findings on an online annotation tool. In the first (unaided) reading session, each reader reviewed all radiographs without DxNodule AI Screen outputs and recorded their findings. The findings were marked as either H (high confidence in detecting nodule), L (Low confidence in detecting nodule), or N (No nodule). After the washout period of one month, a second (aided) reading session was conducted, in which readers reviewed the outputs processed by DxNodule AI Screen along with the original radiographs to record the findings concurrently.

### Statistical analysis

All analysis and computation were performed in python using SciPy statistical library. The comprehensive evaluation of the model performance on the test set included sensitivity, specificity, NPV, accuracy, F1 score, and AUROC. To measure the variability in these values, we used 95% confidence intervals using the empirical bootstrapping method. To compare the AUROC, sensitivity, specificity, accuracy, NPV, and PPV of readers before and after the assistance of DxNodule AI Screen, Wilcoxon signed-rank test was performed on three independent samples of reader annotations. A p-value of less than 0.05 was considered statistically significant. To assess the agreement in nodule identification between the reference standard and DxNodule AI Screen, Bland-Altman (B-A) analysis was performed.

## RESULTS

The findings were considered suggestive of the pulmonary nodule(s) if they had a size between 5-30 mm. These nodules were seen individually or in a cluster in the right and/or the left lung field. These were calcified/non-calcified and were distributed in the upper, mid, or lower zones on the right or left side of the lung field.

As reviewed by the readers who established the ground truth, 103 out of 308 chest radiographs were positive for pulmonary nodules, while the rest (205 radiographs) were negative for pulmonary nodules. Patient information for the number of chest radiographs and number of nodules per chest radiograph is represented in Table 1.

**Table 1:** Patient information and nodules characteristics from the external test dataset

| Characteristic | Numbers |
| --- | --- |
| **Patient information** |  |
| No. of chest radiographs | 308 |
| No. of chest radiographs with nodules | 103 |
| No. of chest radiographs without nodules | 205 |
| **No. of nodules per chest radiograph** |  |
| One nodule | 85 |
| Two nodules | 11 |
| Three nodules | 3 |
| Four nodules | 2 |
| Five nodules | 2 |
| **Radiograph projection** |  |
| Posterior-Anterior (PA) | 220 |
| Anterior-Posterior (AP) | 88 |

### Standalone performance of DxNodule AI Screen

The standalone model performance for detection of pulmonary nodules included 80 true positives, 180 true negatives, 25 false positives, and 23 false negative identifications (Fig. 2). The model had an accuracy of 0.83 (0.80, 0.88) in detecting nodules on the chest radiographs. The model achieved a sensitivity of 0.78 (0.69, 0.85), specificity of 0.88 (0.83, 0.92), and an AUC of 0.905 (0.87, 0.94). Table 2 indicates the performance metrics of the DL model in the detection of lung nodules.

**Figure 2:**
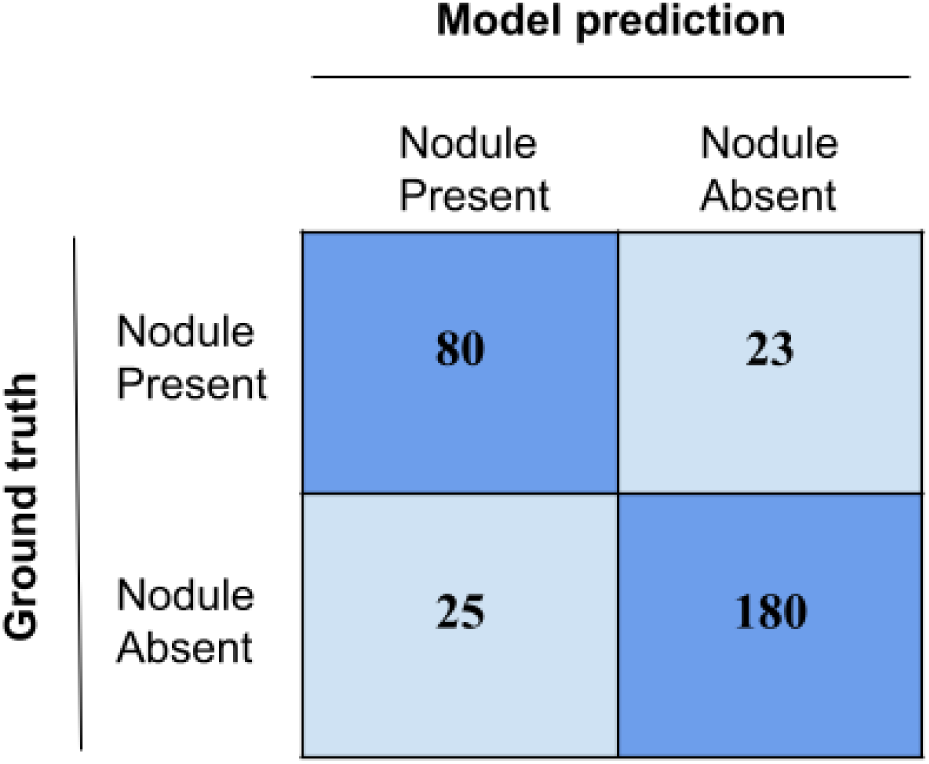
Confusion matrix for AI model on the external test set

**Table 2:** Performance metrics of DxNodule AI Screen

| S.No. | Metrics | Value [95% CI] |
| --- | --- | --- |
| 1 | Accuracy | 0.83 [0.80, 0.88] |
| 2 | Sensitivity | 0.78 [0.69, 0.85] |
| 3 | Specificity | 0.88 [0.83, 0.92] |
| 4 | F1 score | 0.77 [0.70, 0.83] |
| 5 | NPV | 0.89 [0.84, 0.93] |
| 6 | PPV | 0.76 [0.68, 0.84] |
| 7 | AUC | 0.905 [0.87, 0.94] |

### Comparison between the reference standard and DxNodule AI Screen

The Bland-Altman (B-A) analysis was used to demonstrate agreement between the reference standard (ground truth) and DxNodule AI Screen in identifying positive nodules. A mean difference value close to zero represents that DxNodule AI Screen performed well in comparison to the reference standard. Our analysis demonstrated a mean agreement difference of 0.036 (95% CI: -1.307, 1.379) for the test dataset (Fig. 3).

**Figure 3:**
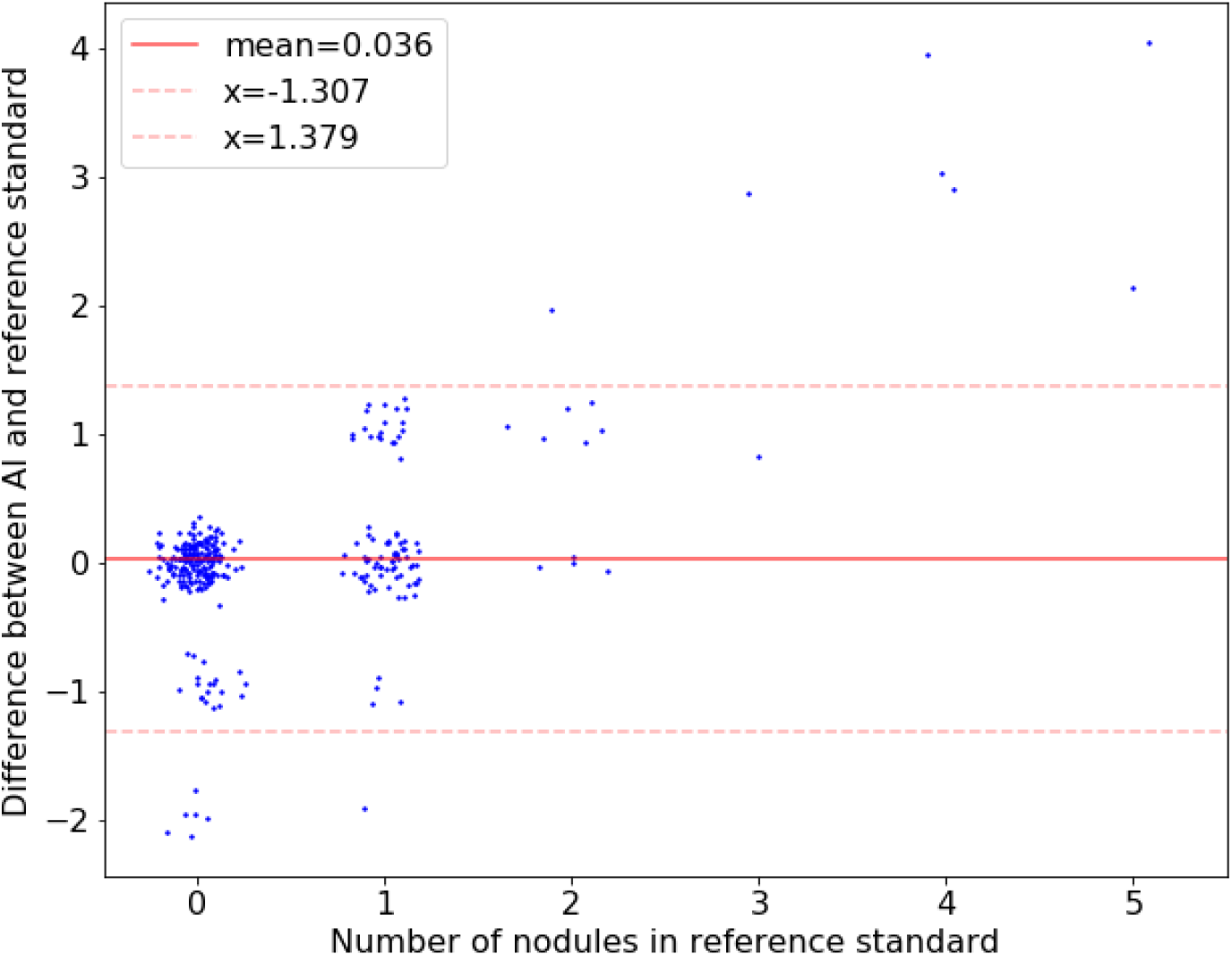
B–A plot illustrating the agreement test for nodule identification between the reference standard and the DxNodule AI Screen. The plots elucidate the difference in the number of nodules identified by the expert ground truther readers (reference standard) and the DxNodule AI Screen (y-axis) over the number of nodules in the reference standard (x-axis). The value of the mean difference (red solid line) and the upper and lower limits of 95% agreement (red dotted lines) are shown. The density of the dots represents the frequency the data appeared on the specific X–Y axis area.

### Observer performance test

All 11 readers assessed radiographs with and without the aid of DxNodule AI Screen. The mean performance of unaided readers for detection of pulmonary nodules included 178.58 true negative, 71.42 true positive, 31.58 false negative and 26.42 false positive identifications. The mean performance of the readers improved with AI-aided interpretation with a decrease in false negative (24.64) and false positive (22.18) identifications and an increase in true negative (182.82) and true positive (78.36) identifications compared with unaided interpretation (Table 3).

**Table 3:** Distribution of lung nodules with DxNodule AI Screen-unaided and DxNodule AI Screen-aided sessions

| Reader | Unaided interpretations |  |  |  | Aided interpretations |  |  |  |
| --- | --- | --- | --- | --- | --- | --- | --- | --- |
|  | TP | TN | FP | FN | TP | TN | FP | FN |
| **R1** | 90 | 143 | 62 | 13 | 88 | 170 | 35 | 15 |
| **R2** | 24 | 184 | 21 | 79 | 84 | 185 | 20 | 19 |
| **R3** | 84 | 140 | 65 | 19 | 89 | 170 | 35 | 14 |
| **R4** | 83 | 191 | 14 | 20 | 77 | 192 | 13 | 26 |
| **R5** | 81 | 190 | 15 | 22 | 70 | 200 | 5 | 33 |
| **R6** | 53 | 196 | 9 | 50 | 56 | 202 | 3 | 47 |
| **R7** | 73 | 200 | 5 | 30 | 81 | 195 | 10 | 22 |
| **R8** | 95 | 126 | 79 | 8 | 97 | 129 | 76 | 6 |
| **R9** | 72 | 193 | 12 | 31 | 81 | 194 | 11 | 22 |
| **R10** | 72 | 199 | 6 | 31 | 59 | 202 | 3 | 44 |
| **R11** | 50 | 201 | 4 | 53 | 80 | 172 | 33 | 23 |
| **Mean**<br>**(SD)** | 71.42<br>(20.02) | 178.58<br>(26.50) | 26.42<br>(26.5) | 31.58<br>(20.02) | 78.36<br>(12.44) | 182.82<br>(21.76) | 22.18<br>(21.76) | 24.64<br>(12.44) |

With the aid of DxNodule AI Screen, the mean specificity, balanced accuracy, PPV and NPV across the 11 readers improved with statistical significance (p<0.05) compared with unaided interpretation. The mean sensitivity and specificity of the readers improved from 0.69 (0.55, 0.82) and 0.87 (0.78, 0.96) in the unaided session to 0.76 (0.68, 0.84) and 0.89 (0.82, 0.96), respectively in the aided session. The mean accuracy, PPV, and NPV of the readers improved from 0.78 (0.72, 0.84), 0.77 (0.65, 0.88), and 0.86 (0.81, 0.90) in the unaided session to 0.83 (0.80, 0.85), 0.82 (0.73, 0.9), and 0.89 (0.86, 0.92), respectively in the aided session (Table 4).

**Table 4:** Sensitivity, Specificity, Accuracy, NPV, and PPV of unaided and DxNodule AI Screen-aided interpretation modes for pulmonary nodule detection

| Reader | Unaided interpretations |  |  |  |  | Aided interpretations |  |  |  |  |
| --- | --- | --- | --- | --- | --- | --- | --- | --- | --- | --- |
|  | Sensitivity | Specificity | Accuracy | NPV | PPV | Sensitivity | Specificity | Accuracy | NPV | PPV |
| R1 | 0.874 | 0.698 | 0.786 | 0.917 | 0.592 | 0.854 | 0.829 | 0.842 | 0.919 | 0.715 |
| R2 | 0.233 | 0.898 | 0.565 | 0.700 | 0.533 | 0.816 | 0.902 | 0.859 | 0.907 | 0.808 |
| R3 | 0.816 | 0.683 | 0.749 | 0.881 | 0.564 | 0.864 | 0.829 | 0.847 | 0.924 | 0.718 |
| R4 | 0.806 | 0.932 | 0.869 | 0.905 | 0.856 | 0.748 | 0.937 | 0.842 | 0.881 | 0.856 |
| R5 | 0.786 | 0.927 | 0.857 | 0.896 | 0.844 | 0.680 | 0.976 | 0.828 | 0.858 | 0.933 |
| R6 | 0.515 | 0.956 | 0.735 | 0.797 | 0.855 | 0.544 | 0.985 | 0.765 | 0.811 | 0.949 |
| R7 | 0.709 | 0.976 | 0.842 | 0.870 | 0.936 | 0.786 | 0.951 | 0.869 | 0.899 | 0.890 |
| R8 | 0.922 | 0.615 | 0.768 | 0.940 | 0.546 | 0.942 | 0.629 | 0.786 | 0.956 | 0.561 |
| R9 | 0.699 | 0.941 | 0.820 | 0.862 | 0.857 | 0.786 | 0.946 | 0.866 | 0.898 | 0.880 |
| R10 | 0.699 | 0.971 | 0.835 | 0.865 | 0.923 | 0.573 | 0.985 | 0.779 | 0.821 | 0.952 |
| R11 | 0.485 | 0.980 | 0.733 | 0.791 | 0.926 | 0.777 | 0.839 | 0.808 | 0.882 | 0.708 |
| Mean<br>(95% CI) | 0.69<br>(0.55, 0.82) | 0.87<br>(0.78, 0.96) | 0.78<br>(0.72,0.84) | 0.857<br>(0.81,0.90) | 0.77<br>(0.65, 0.88) | 0.76<br>(0.68, 0.84) | 0.89<br>(0.82, 0.96) | 0.83<br>(0.80,0.85) | 0.89<br>(0.86, 0.92) | 0.82<br>(0.73, 0.90) |
| p-value |  |  |  |  |  | 0.208 | 0.021 | 0.009 | 0.054 | 0.008 |

There was a significant difference in the performance of radiologists aided by DxNodule AI Screen and those not aided by DxNodule AI Screen. The diagnostic performance of the unaided readers was compared to aided readers and represented as the AUROC curve. Each reader marked an image as either N (No nodule), L (Low confidence in detecting nodule), or H (high confidence in detecting nodule). These three labels were converted to scores: 0, 0.5, and 1 respectively. Using these scores, ROC curves for each reader in both the sessions (aided and unaided) were generated. To get the average ROC curve for all readers, the TPR was averaged for every FPR according to the method described by Chen *et al*. ^15^. The average AUC of the readers improved from 0.798 [0.74, 0.86] to 0.846 [0.82, 0.88] when aided by DxNodule AI Screen (p-value = 0.013). Standalone DxNodule AI Screen achieved an AUC of 0.905 [0.87,0.94] in identifying pulmonary nodules in the test dataset (Fig. 4).

**Figure 4:**
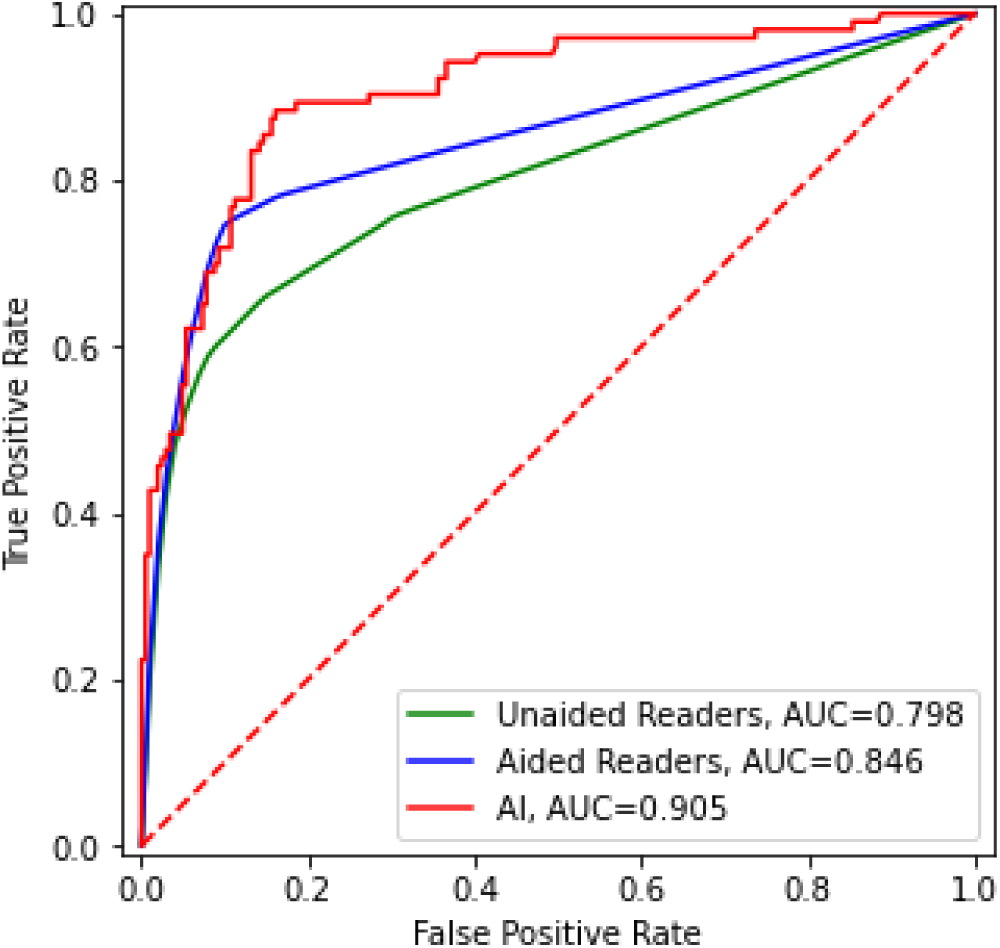
AUROC curves depicting the performance of standalone DxNodule AI Screen (red), DxNodule AI Screen-unaided readers (green), and DxNodule AI Screen-aided readers (blue).

### Lung nodule detection with the aid of DxNodule AI Screen

DxNodule AI Screen assisted the radiologists in identifying nodules that were otherwise missed due to a wide variety of factors like overlying ribs or scapula shadow, small size of nodules, etc. (Fig. 5). DxNodule AI Screen not only helped readers identify radiographs with nodules, but it also helped readers locate nodules more accurately. Fig. 6 represents a clinical case wherein in the absence of DxNodule AI Screen, only 1 reader could mark both the nodules correctly, 7 readers marked only one nodule, 2 readers marked nodules at more than two locations, and 1 reader marked the radiograph as negative. With the help of DxNodule AI Screen, 4 readers could correctly locate both the nodules in the radiograph and there were no false-positive findings.

**Figure 5:**
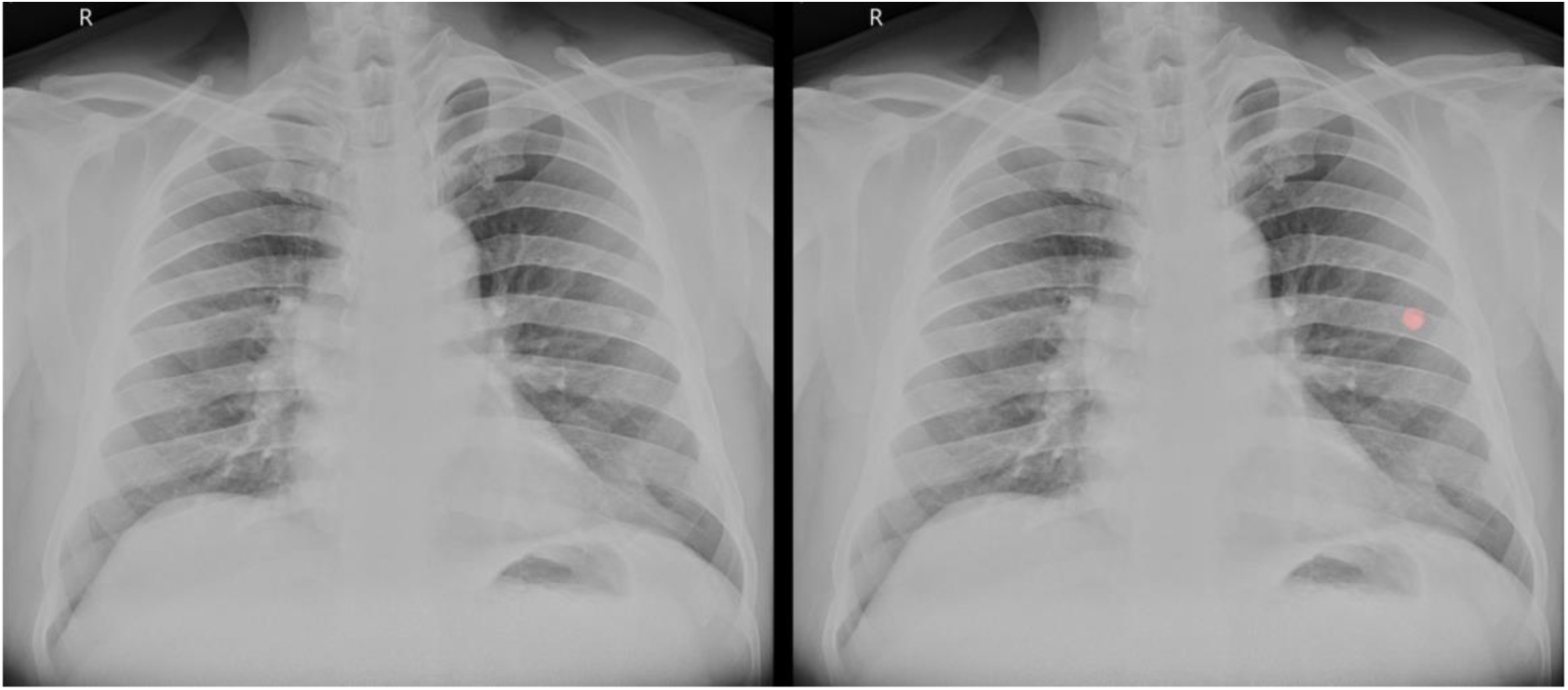
Lung nodule overshadowed by the posterior end of the seventh rib in the left lung. Only 1 out of 11 readers detected this without DxNodule AI Screen. With DxNodule AI Screen, 8 were able to detect.

**Figure 6:**
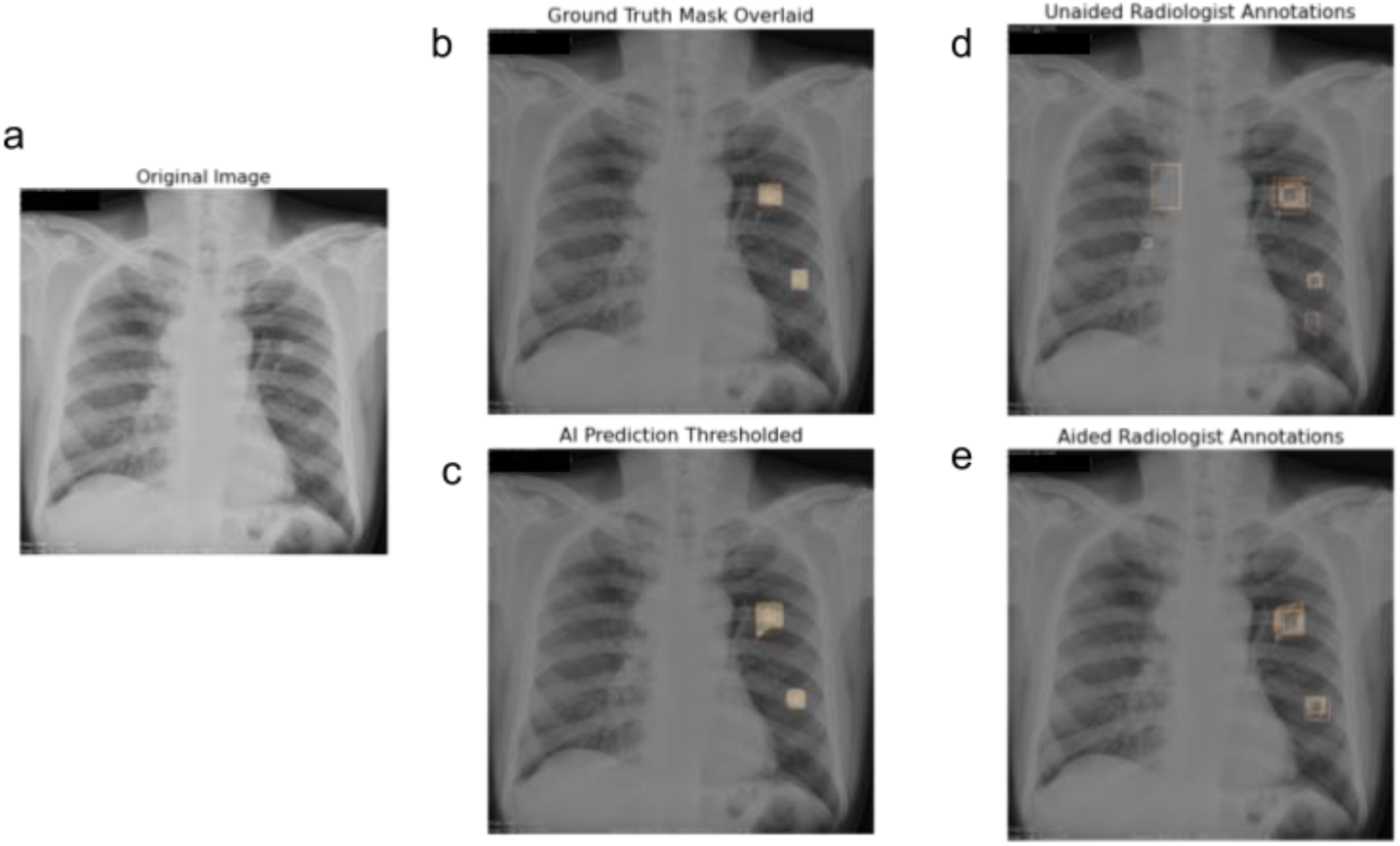
**(a)** Original chest radiograph with two nodules in the left lung. **(b)** Ground truth masks indicate the location of nodules in the chest radiograph. **(c)** Location of nodules as predicted by the DxNodule AI Screen. **(d)** Without the AI assistance, only one reader properly identified both nodules, while seven radiologists identified only one nodule, two readers identified nodules at more than two places (false positive), and one reader marked the radiograph as negative (false negative). **(e)** When aided by AI, four readers could identify both nodules (true positive), six readers marked one nodule as positive, and one radiologist marked the radiograph as negative. There were no false-positive findings.

## DISCUSSION

Our study demonstrated that AI could detect pulmonary nodules on chest radiographs with better AUC than that of the human readers and that it improved readers’ performance when used as an aid. Our multi-reader diagnostic study demonstrated a significant improvement in specificity, accuracy, and AUC in the detection of pulmonary nodules on chest radiographs. The AUCs reported in our study surpass those reported in previous studies. In a study by Homayounieh *et al*., 100 chest radiographs from multiple institutes were evaluated by 9 readers resulting in an increase in mean AUC from 71.7% (unaided read) to 77.2% (aided read) ^16^. Jang et al. reported an increase in the overall AUC of readers from 67% to 76% when using their DLAD system ^17^. Although our study was single-centric, we used a dataset of 308 radiographs to evaluate the performance of human readers with or without the aid of DxNodule AI Screen. We observed an increase in mean AUC from 79.8% (without AI) to 84.6% (with AI), which was superior to the mean AUCs reported by Homayounieh *et al*. and Jang *et al*.

The AI model used in this study demonstrated improved sensitivity and specificity in detecting pulmonary nodules on chest radiographs as compared to unaided interpretation. The mean sensitivity and specificity of the readers improved from 0.69 [0.55, 0.82] and 0.87 [0.78, 0.96] in the unaided session to 0.76 [0.68, 0.84] and 0.89 [0.82, 0.96], respectively in the aided session. Moreover, there was a remarkable decrease in the number of false positives (from 26.42 to 22.18) and false negatives (from 31.58 to 24.64) when DxNodule AI Screen was used as an aid. This may reduce the number of unnecessary follow-up procedures, which can pose a radiation risk.

Considering the high miss rate and importance of identifying pulmonary nodules on chest radiographs, AI algorithms like the one tested in our study can play an important role in improving the quality and diagnostic value of radiographs in patients with pulmonary nodules. Our findings suggest that using DxNodule AI Screen to interpret chest radiographs can help minimize the incidence of missed nodules or false location of nodules and allow for earlier diagnosis of lung cancer without the use of unnecessary chest CT examinations in healthy individuals. These findings imply that the AI system can help in reducing human error and improving the detection accuracy of nodule chest radiographs. Since lung cancer is the primary cause of cancer-related mortality ^18,19^, earlier detection with AI aid may reduce lung cancer mortality in the population.

Our study had some limitations. First, the study sample size was limited to 308 images. To overcome the limitation of the small sample size, we had a total of 6776 reads (11 readers × 308 images × 2 reading sessions) with the focus on only 1 pathological finding (i.e., nodules) to enhance the measurement reliability of readers’ improvement with AI and comparing readers’ mean AUC with and without AI. To avoid overanalysis of a limited data set, we did not evaluate reader performance in detecting other pathologies. Second, our AI model utilizes imaging data from each radiograph to arrive at results and it does not utilize specific patient information, such as age, smoking history, lung disorders, and other related morbidities, which can aid with lung cancer screening and management ^20^. Third, since this was a single-centre retrospective study, it did not accurately represent the real-world situation. Further research is required to determine the applicability of DxNodule AI Screen in a prospective multi-institutional study.

In conclusion, we tested a deep learning-based AI model that outperformed experienced human readers in detecting nodules on chest radiographs. Readers demonstrated improved performance for nodule detection when this model was used as an aid.

## Data Availability

All data produced in the present work are contained in the manuscript

## Acknowledgments

We would like to thank Dr. Reetika Kapoor, Dr. Karthik M., Dr. Shreeya Goel, Dr. Tejvir Singh, and Dr. Suhas M. from Dr. DY Patil Medical College, Hospital and Research Center, Pune, India for their participation in the observer performance study.

## Notes

### Competing Interest Statement

Amit Kharat is a Professor at Dr. DY Patil Medical College, Pune, and is also a co-founder of DeepTek Medical Imaging Pvt Ltd. Vinay Duddalwar is a Professor of Radiology at Keck School of Medicine, USC, USA, and is also on the advisory board of DeepTek Medical Imaging Pvt Ltd. He is a consultant to Radmetrix Inc, Cohere Inc, and Westat Inc.

### Funding Statement

This study did not receive any funding

### Author Declarations

Ethics committee of Dr. D.Y. Patil hospital and research center gave ethical approval for this work

### Summary of Updates

This version of the manuscript has been revised to update the author list

